# Translation of miRNA blood-based discovery to molecular testing for clinical diagnosis of endometriosis

**DOI:** 10.1101/2025.07.23.25332110

**Authors:** Yanqin Yu, Wing Hing Wong, Yao Hu, Yirong Shen, Xiaochun Xu, Shen Lu, Libo Zhu, Xinmei Zhang, Farideh Z Bischoff

## Abstract

Endometriosis is a common yet often underdiagnosed condition, partly due to the lack of reliable diagnostics. This study examines the clinical feasibility of a blood-based, miRNA-driven test to diagnose endometriosis and address the challenges of translating next-generation sequencing (NGS) findings into clinical use. Serum from 20 patients and 20 controls underwent miRNA sequencing to identify diagnostic biomarkers. A machine learning model built on all NGS-based differentially expressed miRNA biomarkers achieved ≥90% accuracy. Validation by qPCR confirmed some but not all findings, underscoring the difficulty of adapting NGS discoveries for routine diagnostics. Nonetheless, serum miRNA biomarkers show strong promise for non-invasive endometriosis detection, with further optimization needed for clinical translation.

## Introduction

Endometriosis is a chronic and often debilitating condition that affects approximately 5-10% of women and adolescents within the reproductive age range of 15-49 years (1–3). However, among women experiencing infertility, the prevalence of endometriosis can rise significantly, affecting approximately 50% of this population. The disease can manifest as early as the first menstrual period and persist until menopause, causing a wide range of symptoms that significantly impact quality of life. Notably, between 50% and 80% of women who suffer from chronic pelvic pain are diagnosed with endometriosis, highlighting its strong association with this symptom (1–3).

Despite its prevalence and impact, endometriosis remains underdiagnosed and poorly understood. Early detection and timely intervention are crucial for improving patient outcomes, yet diagnosis is frequently delayed by an average of 6 to 11 years (4, 5). This delay is attributed to several factors, including the unknown etiology of the disease, the nonspecific and varied nature of its symptoms, and the limitations of current diagnostic methods. Symptoms such as dysmenorrhea, chronic pelvic pain, dyspareunia, and gastrointestinal disturbances can often mimic other gynecologic or gastrointestinal conditions, leading to misdiagnosis or delayed recognition. This variability, coupled with the lack of reliable diagnostic tools, poses significant challenges for both clinicians and researchers (6, 7).

The current “Gold Standard” diagnostic endometriosis test involves an invasive laparoscopic surgical procedure, which carries risks of infection, bleeding, and damage to surrounding tissues (8–10). The procedure is only as reliable as the operator’s surgical experience in locating and identifying endometriosis lesions. As a result, its diagnostic accuracy varies greatly, with some studies reporting sensitivity and specificity below 70% (10, 11). For instance, a study in Canada found that while laparoscopic visualization had a sensitivity of 90%, its specificity was only 40% (12). Another study reported a sensitivity of 69% and a specificity of 83% for laparoscopy in diagnosing endometriosis (13). These findings highlight that while laparoscopy is a valuable diagnostic tool, it is not infallible. Besides relying heavily on the operator’s experience, the heterogeneous appearance of endometriotic lesions precluded a standardized visual assessment, and the presence of deep infiltrating lesions may be missed during the procedure (10, 14). In addition, data suggest that some women undergoing the procedure do not have the condition and are therefore unnecessarily exposed to surgical risks. Further diagnostic advances may be expected from ever-evolving and indeed less invasive imaging technologies, however as yet this has not achieved the gold standard status.

Given these limitations, there is an increasing interest in developing non-invasive diagnostic methods for endometriosis (15–17). A growing body of evidence highlights the significant role of miRNA expression changes in the pathogenesis and progression of various diseases, including endometriosis. Numerous research groups have focused on the regulatory role of miRNAs in key genes associated with endometriosis and have explored their potential as diagnostic biomarkers. For example, studies have identified *miR-200* and *miR-17-5p* as promising biomarkers in the plasma of endometriosis patients (18, 19), while *miR-135a* has shown diagnostic potential in the saliva of these patients (20). However, conflicting results have also emerged regarding the diagnostic value of miRNAs (21). For example, one study found that serum *miR-199a* was upregulated in individuals with endometriosis (22), while another study reported its downregulation in serum (23), highlighting the challenges in interpreting these findings and developing reliable diagnostic tests. Several factors may contribute to these discrepancies. For instance, the menstrual phase during which samples are collected may influence miRNA expression, as studies have shown that miRNA levels can vary between the proliferative and secretory phases of the menstrual cycle (24, 25). Additionally, variations in study protocols, such as differing inclusion and exclusion criteria, may introduce further inconsistencies, especially considering the heterogeneity of endometriosis as a disease. Despite the numerous challenges associated with standardizing miRNA-based diagnostics, such as biological variability, technical inconsistencies, and the complexity of endometriosis as a disease, considerable progress has been made in developing and implementing diagnostic tools in clinical settings. One notable example is the Endotest developed by Ziwig Inc., a test that utilizes a next-generation sequencing (NGS) panel comprising over 100 microRNAs to evaluate disease status (26, 27). This test has been primarily introduced in European healthcare systems and represents a significant advancement in non-invasive diagnostics for endometriosis. Although the Ziwig Endotest holds substantial potential as a laboratory-developed test (LDT), its broader implementation as a routine *in vitro* diagnostic (IVD) tool remains limited. This is largely due to the high cost associated with NGS technologies and the requirement for advanced bioinformatics infrastructure and expertise (28, 29), which may not be readily available in routine clinical settings. As such, while promising, NGS-based test is currently better suited for use in a centralized reference laboratory rather than widespread clinical deployment in the form of IVD (30).

One major obstacle in translating NGS-derived miRNA biomarkers into clinically actionable PCR-based assays (qPCR or ddPCR) is the challenge of selecting appropriate endogenous controls for data normalization (31, 32). Unlike NGS, which employs global normalization strategies such as reads per kilobase million (RPKM), and transcripts per million (TPM), PCR-based approaches require specific endogenous reference genes for normalization. However, many studies rely on “universal” endogenous controls, such as *GAPDH*, *RNU48*, or *miR-16*, without assessing their stability in the specific disease contexts (31, 32). This oversight can introduce systematic bias, leading to unreliable and non-reproducible biomarker quantification, thereby limiting their clinical utility.

To address these challenges, we conducted a proof-of-concept study integrating unbiased miRNA biomarker discovery via miRNA sequencing (miRNA-seq) with subsequent experimental validation using qPCR. Our primary objective is to present the development and feasibility of a miRNA-based panel of biomarkers that would enable a non-invasive, simple and accurate blood test in diagnosing endometriosis reliably. To control for female-specific physiological processes influenced by hormonal fluctuations, such as those occurring during the menstrual cycle, we limited serum collection to women in the secretory phase. Another key innovation of our study is that we not only identified potential biomarkers but also established endometriosis-specific reference endogenous controls for normalization in clinical diagnostic settings. These newly identified endogenous controls were systematically validated to ensure their stability and suitability for qPCR– and ddPCR-based assays. Experimental validation of both the candidate biomarkers and the newly selected endogenous controls demonstrated strong reproducibility and accuracy, supporting their potential clinical application. This approach represents a critical step toward the development of a standardized, reliable, and clinically translatable miRNA-based diagnostic assay for endometriosis.

## Results

### Differentially expressed miRNAs in endometriosis patients

In the discovery cohort of samples collected during the secretory phases, we identified a total of 85 differentially expressed miRNAs between the patient group and the control group (Figure 1A; Supplementary Table S1). Among these, 55 miRNAs were significantly upregulated in the disease group, while 30 were downregulated, indicating a distinct molecular signature associated with endometriosis. Interestingly, some subjects exhibited higher expression levels of specific miRNAs compared to others within the same group. This variability is likely attributable to the heterogeneous nature of endometriosis, which can manifest differently across individuals in terms of severity, symptom presentation, and molecular profiles. Notably, several miRNAs previously implicated in the pathogenesis and progression of endometriosis were found to be highly dysregulated in the disease group compared to the control group (Figure 1A-B). These included *miR-9-5p* and *miR-21-5p*, both of which have been reported to play critical roles in regulating inflammatory signaling pathways, a key mechanism underlying the development and progression of endometriosis.

**Figure 1:**
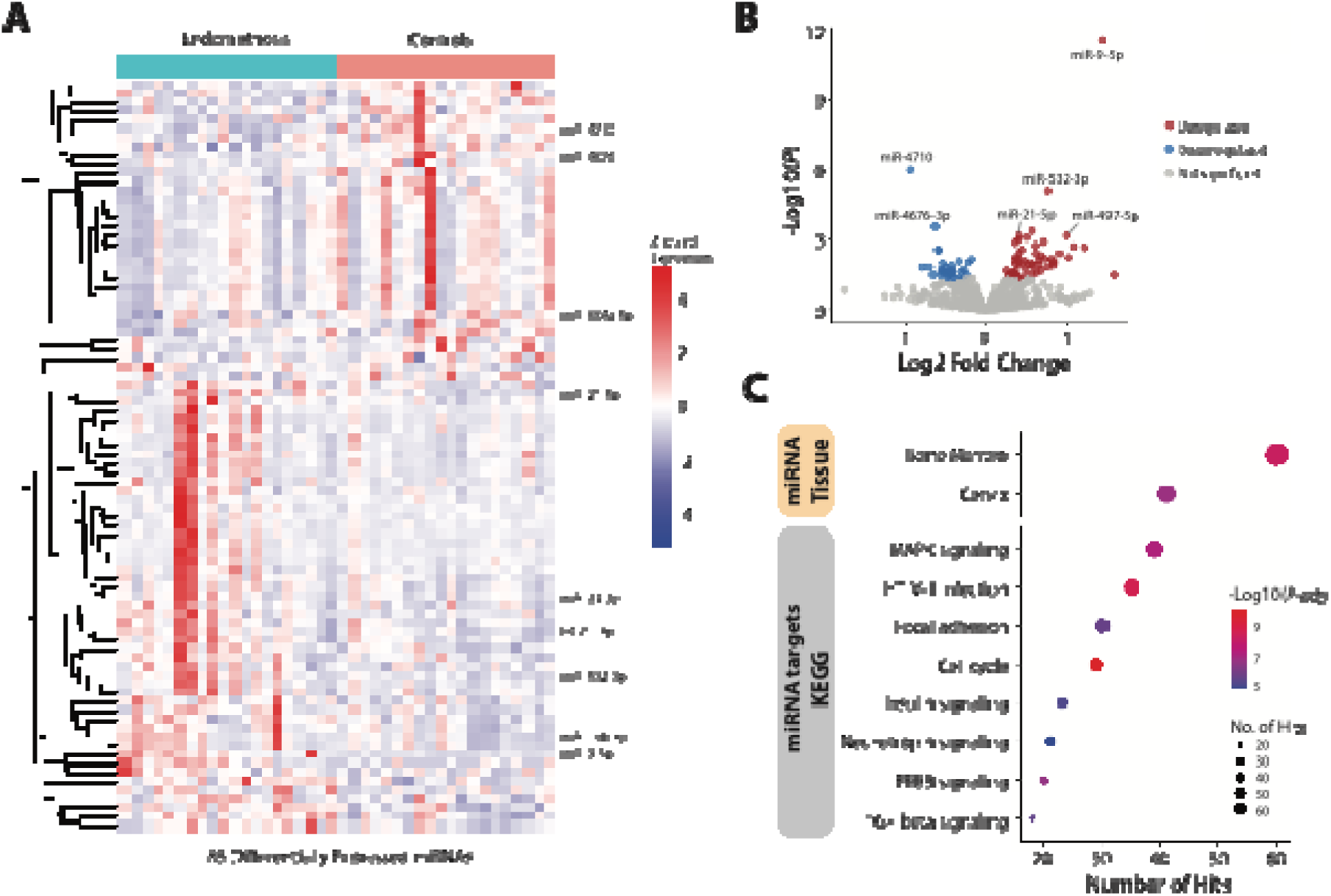
Differential expression analysis of miRNAs in endometriosis patients. (A) Heatmap illustrating the expression profiles of miRNAs that were differentially expressed between endometriosis patients and healthy controls. A total of 85 miRNAs showed significant expression differences. Notably, several of these miRNAs, including miR-9-5p, miR-21-5p, and let-7b-3p, have been previously associated with the pathogenesis of endometriosis and are highlighted on the plot. (B) Volcano plot depicting the distribution of differentially expressed miRNAs based on fold change and statistical significance. (C) Pathway enrichment analysis of the differentially expressed miRNAs. The top panel shows inferred tissue-of-origin patterns, suggesting potential source tissues of the dysregulated miRNAs. The bottom panel presents enriched non-cancer-related KEGG pathways derived from the predicted target genes of these miRNAs, highlighting relevant biological processes potentially involved in endometriosis pathophysiology.

To further investigate the origin and functional relevance of these differentially expressed miRNAs, we performed a hypergeometric over-representation analysis using miRNet (33, 34). This analysis revealed that the majority of these miRNAs predominantly originated from the bone marrow and cervix (Figure 1C). The bone marrow association reflects the serum origin of the miRNA data. The cervix, on the other hand, is relevant as it points to the anatomical regions where endometriosis lesions can be found in proximity, providing confidence that the observed molecular signals are indeed linked to the disease. This suggests that the serum circulating miRNAs may serve as biomarkers for systemic changes associated with endometriosis

Additionally, we conducted a hypergeometric test using the gene targets of the differentially expressed miRNAs against the KEGG database to identify enriched molecular pathways (34). This analysis highlighted that the top non-tumor-specific molecular processes included various signaling pathways and focal adhesion, which are strongly associated with endometriosis (Figure 1C). For instance, focal adhesion is a critical process in the pathogenicity of endometriosis, as it involves cell-matrix interactions that contribute to the attachment, survival, and invasion of endometrial cells outside the uterus (35). Similarly, dysregulation in signaling pathways, such as the MAPK signaling pathway has been observed in endometrial cells, leading to enhanced cell proliferation, endometriosis lesion establishment, and disease persistence (36). Furthermore, MAPK signaling is implicated in pain sensitization, suggesting a role in the chronic pelvic pain experienced by many patients with endometriosis (37).

### Assessment of differentially expressed miRNAs for prediction of endometriosis using machine learning

To evaluate whether the identified differentially expressed miRNAs possess predictive value in distinguishing individuals with endometriosis from control subjects, we implemented and assessed three distinct random forest models, each utilizing a different dataset.

For the first model (Model #1), we constructed a random forest classifier using all 85 differentially expressed miRNAs. To rigorously assess its performance, we employed a 30-fold repeated subsampling cross-validation approach. Model #1 demonstrated strong predictive capability, achieving an overall sensitivity of 0.91, specificity of 0.88, and an area under the curve (AUC) of 0.95 (Figure 2A). To further refine the model and enhance interpretability, we explored feature selection based on importance scores derived from the initial model. Specifically, we developed two additional models with reduced feature sets: Model #2, which utilized the top 40 most informative miRNAs, and Model #3, which incorporated only the top 20 (Supplementary Table S2). These refined models were constructed using the same random forest framework and evaluated with the identical repeated subsampling cross-validation procedure. By focusing on the most predictive miRNAs, we aimed to improve classification accuracy while eliminating noise introduced by less informative features.

**Figure 2:**
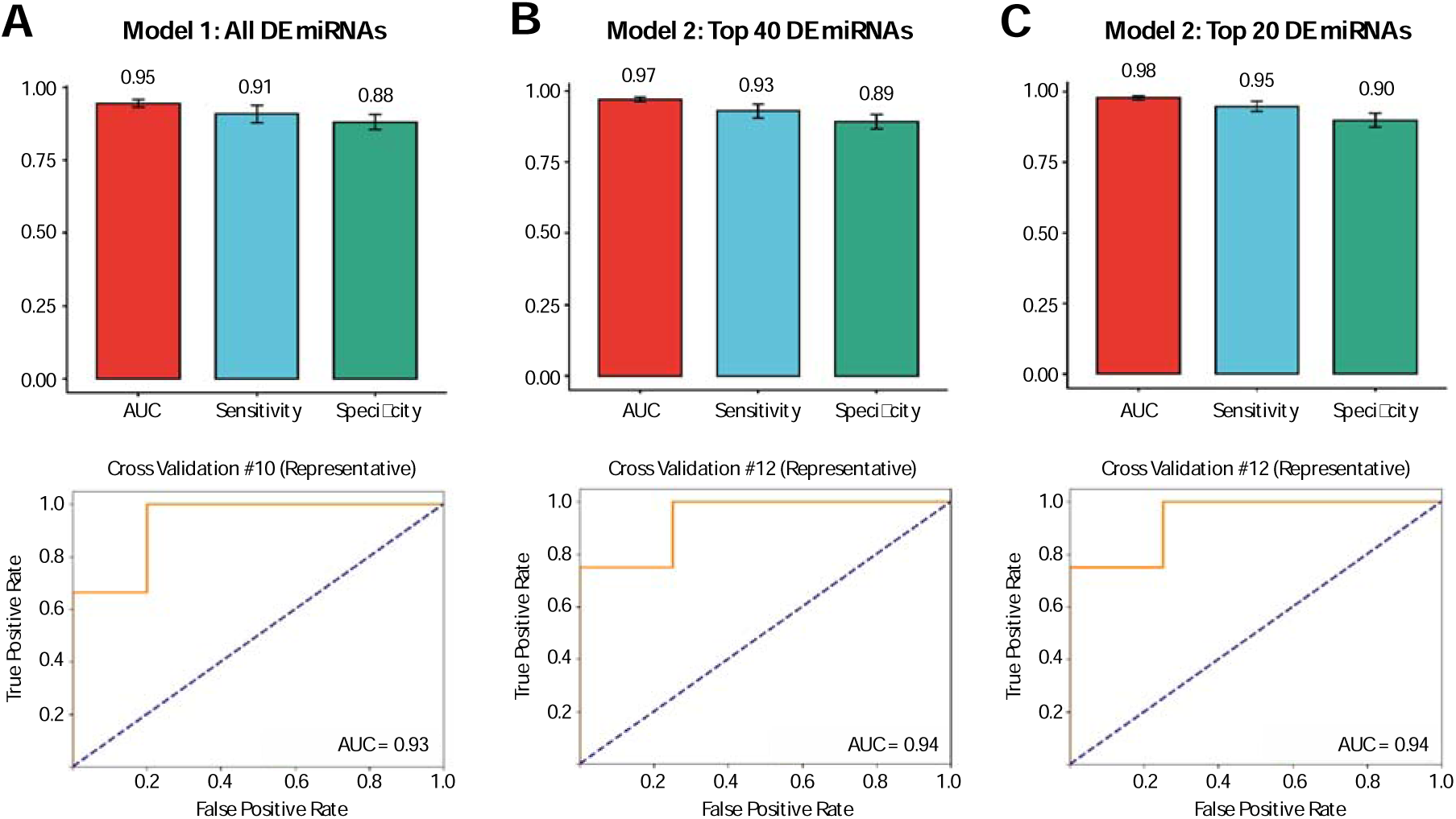
Diagnostic performance of NGS-identified differentially expressed miRNAs using machine learning. The diagnostic utility of differentially expressed miRNAs was assessed through machine-learning analysis. Performance metrics represent the mean of 30 iterations of repeated subsampling cross-validation, with error bars indicating variability across iterations. (A) Using the full set of differentially expressed miRNAs, the model achieved an AUC of 0.95, with a sensitivity of 0.91 and specificity of 0.88. A representative ROC curve from iteration #10 is shown. (B) Limiting the model to the top 40 most informative miRNAs improved performance, yielding an AUC of 0.97, sensitivity of 0.93, and specificity of 0.89. The ROC curve displayed corresponds to iteration #12. (C) Further refinement using the top 20 most informative miRNAs resulted in the highest diagnostic performance, with an AUC of 0.98, sensitivity of 0.95, and specificity of 0.90. The representative ROC curve is also from iteration #12.

The results indicated that both reduced-feature models exhibited slightly improved performance compared to Model #1. Model #2, incorporating the top 40 miRNAs, achieved a sensitivity of 0.93, a specificity of 0.89, and an AUC of 0.97 (Figure 2B). Model #3, which was restricted to the top 20 miRNAs, further enhanced predictive accuracy, yielding a sensitivity of 0.95, a specificity of 0.90, and an AUC of 0.98 (Figure 2C). These findings suggest that a substantial proportion of the differentially expressed miRNAs may not meaningfully contribute to disease classification and instead introduce noise into the predictive model. The improved performance of the reduced-feature models underscores the value of feature selection in enhancing both the robustness and interpretability of machine learning-based biomarker discovery in endometriosis.

### Selection of miRNA biomarkers and endogenous controls for non-NGS clinical diagnostics

The translation of NGS discovery findings into actionable clinical diagnostics is essential for improving patient care. While NGS provides comprehensive molecular insights, clinical diagnostic applications often rely on qPCR due to their faster turnaround time, lower costs, and suitability for IVD implementation without requiring the complex bioinformatics infrastructure necessary for NGS analysis (28–30). Given these advantages, we explored the feasibility of translating our NGS-based findings into a qPCR-based diagnostic assay.

One of the primary considerations in this transition is the limit of detection of qPCR. The 85 differentially expressed miRNAs identified in our NGS analysis exhibit a wide range of expression levels across samples, spanning from as few as 10 normalized read counts (e.g., *miR-4710*) to over 1,000,000 (e.g., *miR-21-5p*) (Supplementary Table S1). Based on initial qPCR assessments, we determined that reliable detection in qPCR is achieved for miRNAs with an average NGS normalized read count exceeding 500. Applying this threshold, 38 of the differentially expressed miRNAs fell below the cutoff and were deemed unreliable for qPCR-based detection (Figure 3A). This left 47 differentially expressed miRNAs that met the detection threshold for further investigation (Supplementary Table S3).

**Figure 3:**
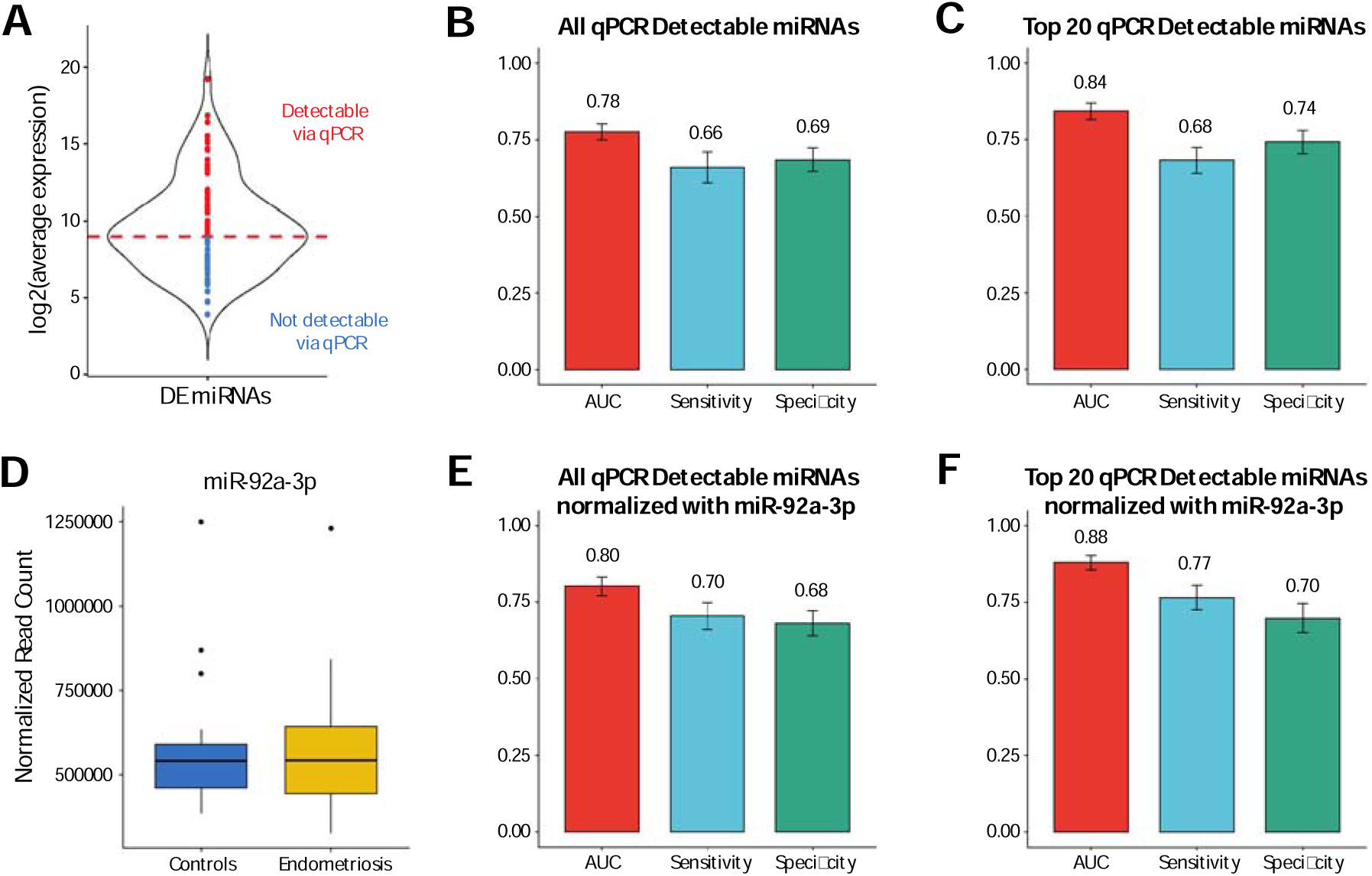
Diagnostic performance of differentially expressed miRNAs reliably quantifiable by qPCR, assessed using machine learning. Performance metrics represent mean of 30 iterations of repeated subsampling cross-validation, with error bars indicating variability across iterations. (A) Violin plot illustrating the expression distribution of differentially expressed miRNAs based on normalized NGS read counts. miRNAs shown in blue represent those with expression levels too low for reliable qPCR detection, while those in red indicate miRNAs with sufficient expression for reliable quantification by qPCR. The dotted red line marks the expression threshold of 500 normalized reads, used to distinguish between the two groups. (B) Predictive performance of a machine-learning model using all 47 miRNAs deemed reliably detectable by qPCR, resulting in an average AUC of 0.78, with a sensitivity of 0.66 and specificity of 0.69. (C) Model performance using the top 20 most informative miRNAs from the qPCR-detectable set, showing a modest improvement with an average AUC of 0.84, sensitivity of 0.68, and specificity of 0.74. (D) Boxplot of miR-92a-3p, a potential endogenous control candidate in this disease setting, demonstrating consistent expression with minimal variability between patient and control groups. The box’s lower and upper hinges correspond to the 25th and 75th percentiles, respectively, with the median indicated by the line inside the box. The whiskers extend to the most extreme data points within 1.5 times the interquartile range below the 25th percentile and above the 75th percentile. (E) Predictive performance using all 47 reliably detectable miRNAs after in silico normalization against miR-92a-3p, yielding an average AUC of 0.80, with sensitivity of 0.70 and specificity of 0.68. (F) Model performance using the top 20 most informative miRNAs following in silico normalization with miR-92a-3p, showing further improvement with an average AUC of 0.88, sensitivity of 0.77, and specificity of 0.68.

**Figure 4:**
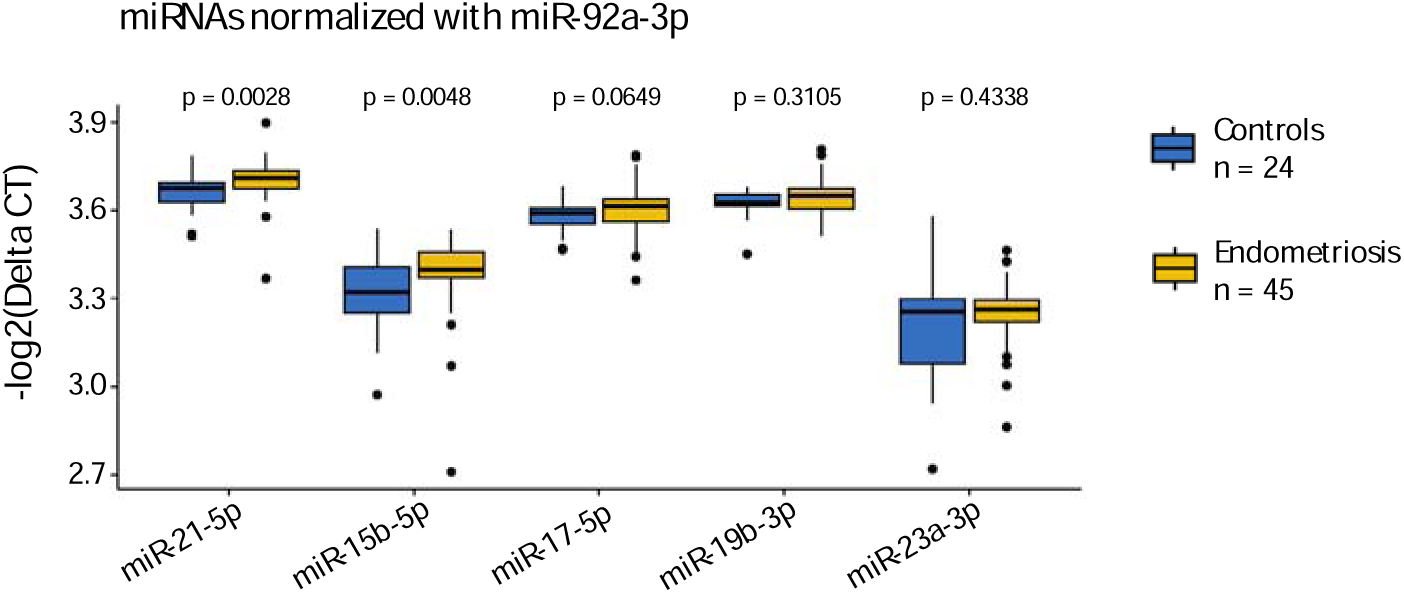
Boxplot of qPCR performance for selected differentially expressed miRNAs. All miRNAs were normalized against miR-92a-3p using qPCR. Of the five miRNAs tested, two (miR-21-5p and miR-15b-5p) showed a significant difference in expression between patients and controls, replicating the direction of effect observed in the NGS discovery dataset. miR-17-5p exhibited a concordant trend, with a p-value of 0.065, approaching statistical significance. The remaining two miRNAs, miR-19b-3p and miR-23a-3p, did not show a statistically significant difference between patients and controls. The boxplot’s lower and upper hinges represent the 25th and 75th percentiles, respectively, with the median indicated by the line inside the box. The whiskers extend to the most extreme data points within 1.5 times the interquartile range below the 25th percentile and above the 75th percentile.

To assess the predictive potential of these 47 miRNAs in a qPCR setting, we constructed two random forest models. The first model incorporated all 47 differentially expressed miRNAs, while the second utilized a reduced feature set consisting of the 20 most informative miRNAs selected from this pool. Notably, many of the miRNAs identified as top contributors in the previous analysis using all 85 markers—such as *miR-21-5p*, *miR-17-5p*, and *miR-15b-5p*—remained among the top-ranked features, suggesting that key predictive biomarkers identified via NGS may indeed be translatable to a clinical qPCR assay.

Performance evaluation of these models revealed that the first model (using all 47 miRNAs) achieved an AUC of 0.78 (Figure 3B), whereas the second model (using the top 20 miRNAs) exhibited improved performance with an AUC of 0.84 (Figure 3C). As observed previously, reducing the feature set led to a slight increase in performance, likely due to the removal of uninformative or noisy variables. However, it is important to note that these qPCR-based models demonstrated lower overall performance compared to the models trained on all 85 differentially expressed miRNAs in the NGS dataset. This suggests that some of the most informative miRNAs are expressed at low levels in the samples, such as *miR-9-5p*, which was ranked as the most predictive but had an average normalized read count of only ∼350. The inability of qPCR to reliably detect such lowly expressed miRNAs presents a challenge for clinical translation. Addressing this limitation may require optimizing primer designs, adopting ddPCR for enhanced sensitivity, or incorporating target capture methods to improve miRNA detection.

A second critical factor in transitioning to a qPCR-based assay is the selection of a suitable endogenous control or reference marker for data normalization. Inappropriate reference markers can introduce variability, leading to unreliable expression quantification and potentially skewing diagnostic outcomes. To address this, we implemented a bioinformatic pipeline in identifying disease-specific endogenous controls (Ref. Materials and Methods). Briefly, we selected miRNAs that display minimal inter-group variability in expression to be used as endogenous references (Supplementary Table S4). One such candidate, *miR-92a-3p* (Figure 3D), demonstrated consistent expression levels between individuals with endometriosis and control subjects. Notably, *miR-92a-3p* has been validated in prior studies as a reliable endogenous control in blood-based miRNA analyses (38).

To further refine our models, we performed *in silico* normalization of the 47 differentially expressed miRNAs against *miR-92a-3p*, simulating the clinical qPCR diagnostic workflow. Following normalization, we re-evaluated the performance of our random forest models. This approach led to noticeable improvements in predictive accuracy, with the model using all 47 miRNAs achieving an AUC of 0.80, and the reduced model using the top 20 miRNAs attaining an AUC of 0.88 (Figures 3E-F). These findings underscore the importance of proper normalization strategies in enhancing the robustness of qPCR-based diagnostics and further support the potential clinical applicability of our NGS-derived biomarker panel.

### Experimental assessment for diagnostic qPCR assays

To evaluate the potential of translating our NGS discovery findings into a clinically viable qPCR diagnostic assay, we selected five candidate miRNAs — *miR-21-5p*, *miR-15b-5p*, *miR-17-5p*, *miR-19b-3p*, and *miR-23a-3p* — for experimental validation using qPCR. These miRNAs were chosen based on their differential expression patterns observed in the NGS dataset, as well as their biological relevance to endometriosis. In addition, *miR-92a-3p* was included as an endogenous control for normalization, given its demonstrated stability across endometriosis and control samples in both our dataset and prior studies.

For this validation study, we conducted qPCR assays on 69 serum samples, comprising 45 patients with endometriosis and 24 control subjects whose disease status was confirmed via laparoscopic surgery. The goal was to determine whether the expression patterns observed in the NGS discovery dataset could be reliably recapitulated using qPCR, a crucial step in transitioning towards a clinically deployable assay. Analysis of the qPCR results revealed that two of the five selected biomarkers (*miR-21-5p* and *miR-15b-5p*) displayed statistically significant differences in normalized expression between the endometriosis and control groups. These findings are consistent with our NGS discovery dataset, reinforcing their potential utility as diagnostic biomarkers. Additionally, *miR-17-5p* exhibited a discernible trend of differential expression between disease and control samples, as visualized in the boxplot analysis. However, this difference did not reach statistical significance, suggesting that while this marker may hold some biological relevance, additional optimization—such as larger sample sizes or refined qPCR conditions—may be necessary to establish its diagnostic value. In contrast, the remaining two miRNAs, *miR-19b-3p* and *miR-23a-3p*, did not show clear delineation between patients and controls, indicating that their differential expression in the NGS dataset may not translate robustly into a qPCR-based assay. This discrepancy underscores the complexities of biomarker translation and highlights the need for rigorous validation and optimization at the experimental level. Factors such as primer design, amplification efficiency, RNA extraction variability, and technical noise could all contribute to differences between NGS and qPCR results, emphasizing the importance of careful assay development. Notwithstanding, the data suggests that qPCR-based assays could provide meaningful diagnostic utility with proper refinement.

## Discussion

Small RNAs such as miRNAs have been shown to play pivotal roles in numerous physiological and pathological processes, influencing gene regulation in both normal and disease states. Altered miRNA expression profiles have been extensively documented in the plasma and serum of patients with various conditions, including diffuse large B-cell lymphoma, ovarian cancer, and type 2 diabetes (39, 40). These findings underscore the potential of circulating miRNAs as non-invasive biomarkers for disease detection and monitoring. In this study, we demonstrate that miRNAs circulating in serum can potentially serve as reliable biomarkers for the diagnosis of endometriosis. The ability to analyze serum miRNA levels in a standardized manner presents a promising approach in disease detection. The fluctuations in specific circulating miRNAs offer a quantifiable and reproducible means of identifying endometriosis, potentially improving early diagnosis and clinical management.

Using serum miRNAs as diagnostic biomarkers offers several key advantages over conventional diagnostic methods in endometriosis. Currently, the gold standard for endometriosis diagnosis relies on laparoscopy with direct visualization, an invasive surgical procedure. A serum-based miRNA biomarker assay could provide a non-invasive alternative, enabling comprehensive disease assessment without the need for surgery. This is particularly valuable for early detection and for patients who may not have immediate access to specialized surgical evaluation. Secondly, compared to invasive diagnostic procedures, a serum-based miRNA test is significantly more cost-effective. The process involves routine blood collection and standard laboratory processing, making it more accessible for widespread clinical implementation. Additionally, standardizing miRNA detection protocols could facilitate large-scale screening efforts, improving early diagnosis and patient outcomes.

In this study, we investigated serum miRNA expression profiles in individuals with endometriosis and identified a distinct set of circulating miRNAs that may serve as potential biomarkers for disease detection. To minimize variability associated with hormonal changes during the menstrual cycle, particularly those impacting female-specific physiological processes, serum samples were collected exclusively during the secretory phase. Our results add to the growing evidence supporting the use of serum miRNA signatures as non-invasive diagnostic tools for endometriosis (15–26). Nonetheless, despite their promise, several key challenges remain before miRNA-based assays can be translated into clinically reliable diagnostic applications.

One of the primary obstacles lies in the choice of detection platform. While NGS provides a comprehensive assessment of the miRNA landscape, its high cost per sample and dependence on complex bioinformatics infrastructure render it impractical for routine clinical diagnostics and IVD applications. Consequently, there is a need to transition toward more practical methodologies, such as qPCR or ddPCR, which offer lower costs, faster turnaround times, and compatibility with IVD requirements.

Our findings suggest that converting NGS-based miRNA discoveries into clinically applicable assays is achievable, though it necessitates experimental refinement. Despite using a limited qPCR panel with only five miRNA biomarkers, we demonstrated valuable diagnostic potential, albeit with a need for further optimization. This highlights both the promise and the technical challenges of implementing miRNA-based diagnostics in clinical practice.

To improve the reliability and diagnostic accuracy of a qPCR-based test, several key strategies should be considered. (a) Primer Optimization: Refining primer designs to enhance amplification efficiency and specificity, particularly for low-abundance miRNAs. (b) Adopting ddPCR for Enhanced Sensitivity: ddPCR offers improved precision and sensitivity, making it a suitable alternative for detecting low-expressed miRNAs that may be missed by qPCR. (c) Expanding Sample Size: Increasing the number of clinical samples analyzed will improve statistical power and ensure robustness across diverse patient populations. (d) Developing a Multiplex Assay: Creating a multiplexed qPCR panel would allow for the simultaneous detection of multiple miRNA biomarkers, streamlining workflow and improving diagnostic efficiency.

While initial qPCR validation of selected miRNAs shows promise, further refinement is necessary and underway to enhance assay reproducibility and clinical performance. Future studies will focus on validating these biomarkers in larger, independent cohorts, optimizing detection methods, and standardizing protocols to ensure reproducibility across clinical testing laboratories. These efforts will be critical in bridging the gap between high-throughput discovery research and real-world clinical application, ultimately paving the way for a clinically deployable serum miRNA-based diagnostic test for endometriosis.

## Materials and Methods

### Specimen collection

Peripheral blood samples were collected prospectively from women aged 18-49 years who presented with mild to severe symptoms, including pelvic pain and/or menstrual bleeding. The participants were enrolled under an approved institutional review board protocol (IRB-20240110-R) at the Women’s Hospital of Zhejiang University School of Medicine, with informed consent obtained from all individuals. The participants included in this study were collected from March 2024 till December 2024. All participants were clinically suspected of a gynecologic abnormal condition and were scheduled to undergo laparoscopy with histopathological evaluation. To account for potential variations in miRNA expression due to different menstrual cycle phases, blood samples were collected exclusively from women in the secretory phase of their menstrual cycle. The menstrual phase was initially determined by physicians or surgeons based on self-reported cycle days and clinical assessment. To ensure accuracy, this classification was further validated through serum progesterone measurements using the protein assay from Kangrun Biotech Co. Ptd (Guangdong, China), with levels exceeding 1.08 ng/mL serving as a biochemical confirmation of the secretory phase according to the manufacturer’s protocol. This approach minimized the influence of hormonal fluctuations on biomarker expression, thereby enhancing the reliability of our findings. A total of 40 symptomatic women were included in the NGS discovery cohort, with 10 mL of blood drawn into standard red-top blood collection tubes prior to the laparoscopic surgery. Among them, 20 women were confirmed to have endometriosis based on both laparoscopic findings and histopathology (disease group), while the remaining 20 had no evidence of endometriosis and served as the control group. Serum was isolated using a two-step centrifugation protocol. First, samples underwent a low-speed centrifugation at 3000 rpm for 10 minutes at 4°C to remove cellular components. This was then followed by a second high-speed centrifugation at 16,000 g for 10 minutes at 4°C to ensure complete removal of debris and platelets. The isolated serum was then aliquoted and stored at –80°C for subsequent RNA extraction and downstream processing.

### RNA isolation and miRNA sequencing (miRNAseq)

Total RNA was isolated from 300 μL of serum using Norgen RNA extraction kit following the manufacturer’s instructions. Total RNAs were ligated to 3’ adapters by denaturation at 70L for 2 minutes, and then incubated at 16L for over 8 hours using NEB T4 RNA Ligase 2. Following 5’ adapters were incubated with previous product using NEB T4 RNA Ligase 1 at 37L for 60 minutes. Ligated RNA was reverse transcribed in a thermocycler using SuperScript II Reverse Transcriptase from ThermoFisher Inc. under the following conditions: an initial incubation at 50L for 60 minutes, followed by a heat inactivation step at 80L for 10 minutes. Following cDNA synthesis, library preparation was performed using the NEB Phusion High-Fidelity DNA Polymerase, adhering strictly to the manufacturer’s guidelines. The final libraries were then subjected to high-throughput sequencing to profile the miRNA expression by LC Biosciences.

### NGS miRNA differential expression profiling endogenous control selection

The raw FASTQ data obtained from miRNA sequencing underwent preprocessing and analysis using the miRge3 (41) software pipeline. Initially, the sequences were quality-trimmed and adapter sequences were removed using a Cutadapt (42) wrapper integrated within miRge3. The trimmed reads were then aligned to the miRBase (43) reference database using Bowtie (44) optimized for short reads. Following alignment, miRge3 generated a comprehensive count table summarizing the abundance of each miRNA across the samples. This count table served as the input for downstream differential expression analysis using the DESeq2 (45) package in R. DESeq2 was employed to identify miRNAs that exhibited statistically significant differences in expression between the patient and control groups. After identifying the differentially expressed miRNAs, hypergeometric over-representation analyses were conducted using the miRNet platform (33). In this analysis, two distinct queries were performed: first, the differentially expressed miRNAs were compared against the miRNA-tissue origin database in miRNet to infer potential tissue-specific origins or associations of these miRNAs; second, the predicted gene targets of these miRNAs were mapped to the KEGG (Kyoto Encyclopedia of Genes and Genomes) database (34) to identify statistically enriched biological pathways. This dual-level approach enabled the contextualization of the miRNA expression patterns in terms of both tissue relevance and functional pathway involvement.

### Endogenous control selection for in silico normalization and qPCR experimental validation

To bridge the findings from the next-generation sequencing (NGS) discovery cohort into clinically applicable diagnostic tools, we further aimed to identify condition-specific endogenous control miRNAs. These controls are essential for normalizing quantitative PCR (qPCR) or droplet digital PCR (ddPCR) assays, ensuring accurate and reproducible quantification of target miRNA expression. The selection of endogenous controls was guided by stringent criteria. Specifically, we evaluated the expression stability of candidate miRNAs by assessing their dispersion estimates and imposing constraints on log2 fold-change (|log2FC| < 0.02) between the patient and control groups. This ensured that the selected controls exhibited minimal variability across conditions. Additionally, candidates were filtered based on an adjusted p-value threshold (≥ 0.8), ensuring that their expression was not influenced by the experimental conditions or disease state.

### Disease prediction model construction using a random forest classifier

To assess the predictive capability of the differentially expressed miRNAs and determine the extent to which they could accurately classify patients with endometriosis, we constructed a random forest classifier using all the differentially expressed miRNAs identified in the discovery cohort. To ensure a robust evaluation of the model’s performance, we performed 30 iterations of repeated random subsampling cross-validation, where in each iteration, the data was split into an 80:20 ratio for training and testing, respectively. This repeated holdout validation approach allowed us to account for variability in model performance due to random data partitioning and provided a reliable estimate of the model’s predictive accuracy. During model construction, any missing values were imputed using the median of the corresponding feature.

Following the initial model construction, we conducted feature selection to identify the most informative miRNAs for predicting endometriosis. This was achieved by evaluating the feature importance scores generated by the random forest algorithm. Based on these scores, we created two distinct feature sets: one comprising the top 40 most important miRNAs and another consisting of the top 20 most important miRNAs. These reduced feature sets were then used to generate and assess new models using the same repeated random subsampling cross-validation procedure described above. Reducing the feature set is crucial for the assessment of model performance and interpretability. A smaller, more informative subset of miRNAs helps prevent overfitting, ensuring the model generalizes well to new data. Additionally, focusing on the most important miRNAs reduces noise, leading to more reliable predictions. A reduced feature set also facilitates translation into clinical diagnostics where the informative miRNAs can be assayed using qPCR and/or ddPCR instead of NGS.

### RNA extraction and qPCR experimental validation

Total RNA was extracted from 200 μL serum using the miRNeasy Serum/Plasma Advanced Kit from Qiagen, following the manufacturer’s recommended protocol. Subsequently, targeted microRNAs (miRNAs) were reverse transcribed into complementary DNA (cDNA) using the FastKing RT Kit II from TianGen Inc. The reverse transcription reaction was carried out in a thermocycler under the following conditions: an initial incubation at 42C for 15 minutes to facilitate cDNA synthesis, followed by a heat inactivation step at 95C for 1 minute to terminate the reactions. The synthesized cDNA was subjected to quantitative PCR analysis of the five miRNA markers. The PCR reaction mixtures were prepared by miRCURY LNA miRNA SYBR Green PCR Kit from Qiagen. qPCR amplification was performed in QuantStudio qPCR system following: an initial denaturation at 95L for 2 minutes, followed of 40 cycles with denaturation at 95 L for 10 seconds and annealing at 56L for 60 seconds.

### Statistical analyses

Differential expression analysis was performed using DESeq2, which models NGS count data with a negative binomial distribution to assess statistical differences between patients and controls. For machine learning-based predictions, sensitivity, specificity, and AUC were evaluated using Python’s scikit-learn package. Pairwise expression comparisons between patients and controls were assessed using the Wilcoxon ranked-sum test.

### Study approval

The participants were enrolled under an approved institutional review board protocol (IRB-20240110-R) at the Women’s Hospital of Zhejiang University School of Medicine, with informed consent obtained from all individuals.

### Data availability

Sequencing data were deposited into Genome Sequence Archive under the accession number PRJCA039165

## Author Contributions

Y. Yu and W.H. Wong led the research group, analyzed the data and wrote the manuscript. Y. Yu, W.H. Wong, X. Zhang and F. Bischoff conceptualized the study. X. Zhang and L. Zhu provided the samples while S. Lu coordinated sample collection between the hospital and the laboratory. W.H. Wong and Y. Hu performed random forest analysis. Y. Yu, Y. Shen and X. Xu performed molecular experiments.

## Acknowledgments

The authors wish to express their gratitude to Jonathan Zhao and Frank Zhang (both from Heranova Lifesciences) for their valuable input on the study design and contributions to the development of the manuscript.

## References

1. Zondervan KT, et al. Endometriosis. NEJM. 2020;382:1244–1256

2. Bonavina G, Taylor HS. Endometriosis-associated infertility: From pathophysiology to tailored treatment. Front Endocrinol. 2022;13:1020827

3. Parasar P, et al. Endometriosis: Epidemiology, diagnosis and clinical management. Curr Obstet Gynecol Rep. 2017;6(1):34–41

4. Fryer J, et al. Understanding diagnostic delay for endometriosis: A scoping review using the social-ecological framework. Health Care for Women International. 2025;46:335–351

5. Kirk UB, et al. Understanding endometriosis underfunding and its detrimental impact on awareness and research. npj Women’s Health. 2024;2:45

6. Davenport S, et al. Barriers to a timely diagnosis of endometriosis: A qualitative systematic review. Obstet Gynecol. 2023;142:571–583

7. Pascoal E, et al. Strengths and limitations of diagnostic tools for endometriosis and relevance in diagnostic test accuracy research. Ultrasound Obstet Gynecol. 2022;60(3):309–327

8. Leyland N, et al. Endometriosis: Diagnosis and management. J Obstet Gynaecol Can. 2010;32:S1–S32

9. Dunselman GA, et al. ESHRE guideline: Management of women with endometriosis. Hum Reprod. 2014;29:400–412

10. Taylor HS, et al. An evidence-based approach to assessing surgical versus clinical diagnosis of symptomatic endometriosis. Int J Gynecol Obstet. 2018;142:131–142

11. Albee RB Jr, et al. Laparoscopic excision of lesions suggestive of endometriosis or otherwise atypical in appearance: Relationship between visual findings and final histologic diagnosis. J Minim Invasive Gynecol. 2008;15:32–37

12. Gratton S, et al. Diagnosis of endometriosis at laparoscopy: A validation study comparing surgeon visualization with histologic findings. J Obstet Gynaecol Can. 2022;44:135–141

13. Sutton CJ, et al. Prospective, randomized, double-blind, controlled trial of laser laparoscopy in the treatment of pelvic pain associated with minimal, mild, and moderate endometriosis. Fertil Steril. 1994;62:696–700

14. Afors K, et al. Employing laparoscopic surgery for endometriosis. Women’s Health. 2014;10:431–443

15. Vanhie A, et al. Plasma miRNAs as biomarkers for endometriosis. Hum Reprod. 2019;34:1650–1660

16. Avery JC, et al. Noninvasive diagnostic imaging for endometriosis part 1: A systematic review of recent developments in ultrasound, combination imaging, and artificial intelligence. Fertil Steril. 2024;121:164–188

17. Oskouei BS, et al. Non-invasive blood tests for earlier diagnosis and treatment of endometriosis. J Reprod Immun. 2025;169:104521

18. Rekker K, et al. Circulating miR-200-family micro-RNAs have altered plasma levels in patients with endometriosis and vary with blood collection time. Fertil Steril. 2015;104:938–946

19. Papari E, et al. Identification of candidate microRNA markers of endometriosis with the use of next-generation sequencing and quantitative real-time polymerase chain reaction. Fertil Steril. 2020;113:1232–1241

20. Perricos A, et al. Hsa-mir-135a shows potential as a putative diagnostic biomarker in saliva and plasma for endometriosis. Biomolecules. 2022;12:1144

21. Vanhie A, et al. Circulating microRNAs as non-invasive biomarkers in endometriosis diagnosis – a systematic review. Biomedicines. 2024;12:888

22. Wang WT, et al. Circulating microRNAs identified in a genome-wide serum microRNA expression analysis as noninvasive biomarkers for endometriosis. J Clin Endocrinol Metab. 2013;98:281–289

23. Hsu CY, et al. miRNA-199a-5p regulates VEGFA in endometrial mesenchymal stem cells and contributes to the pathogenesis of endometriosis. J Pathol. 2014;232:330–343

24. Petracco R, et al. Evaluation of miR-135a/b expression in endometriosis lesions. Biomedical Reports. 2019;11:181–187

25. Lamon S, et al. The effect of the menstrual cycle on the circulating microRNA pool in human plasma: A pilot study. Hum Reprod. 2022;38:46–56

26. Bendifallah S, et al. Machine learning algorithms as new screening approach for patients with endometriosis. Sci Rep. 2022;12:639

27. Ferrier C, et al. Saliva microRNA signature to diagnose endometriosis: A cost-effectiveness evaluation of the Endotest. BJOG. 2023;130:396–406

28. Zalis M, et al. Next-generation sequencing impact on cancer care: Applications, challenges, and future directions. Frontiers in Genetics. 2024;15:1420190

29. Desai K, et al. Real-world trends in costs of next-generation sequencing (NGS) testing in U.S. setting. Journal of Clinical Oncology. 2021;39:15_suppl

30. Karlovich CA, Williams PM. Clinical applications of next-generation sequencing in precision oncology. Cancer J. 2019:25:264–271

31. Chapman JR, Waldenstrom J. With reference to reference genes: A systematic review of endogenous controls in gene expression studies. PLoS ONE. 2015;10(11):e0141853

32. Taylor SC, et al. The ultimate qPCR experiment: Producing publication quality, reproducible data the first time. Trends in Biotechnology. 2019;37(7):761–774

33. Chang L, et al. miRNet 2.0: Network-based visual analytics for miRNA functional analysis and systems biology. Nucleic Acids Research. 2020;48:W244–W251

34. Kanehisa M, Goto S. KEGG: Kyoto encyclopedia of genes and genomes. Nucleic Acids Research. 2000;28(1):27–30

35. Nagai T, et al. Focal adhesion kinase-mediated sequences, including cell adhesion, inflammatory response, and fibrosis, as a therapeutic target in endometriosis. Reproductive Sciences. 2020;27:1400–1410

36. Bora G, Yaba A. The role of mitogen-activated protein kinase signaling pathway in endometriosis. J Obstet Gynaecol Res. 2021;47(5):1610–1623

37. Uimari O, et al. Genome-wide genetic analyses highlight mitogen-activated protein kinase (MAPK) signaling in the pathogenesis of endometriosis. Human Reproduction. 2017;32(4):780–793

38. Solayman MHM, et al. Identification of suitable endogenous normalizers for qRT-PCR analysis of plasma microRNA expression in essential hypertension. Mol Biotechnol. 2016;58(3):179–187

39. Leva GD, Croce CM. miRNA profiling of cancer. Curr Opin Genet Dev. 2013;23(1):3–11

40. Vasu S, et al. MicroRNA signatures as future biomarkers for diagnosis of diabetes states. Cells. 2019;8:1533

41. Patil AH, Halushka MK. miRge3.0: A comprehensive microRNA and tRF sequencing analysis pipeline. NAR Genomics and Bioinformatics. 2021;3(3):lqab068

42. Marcel M. Cutadapt removes adapter sequences from high-throughput sequencing reads. EMBnet.journal. 2011;17(1):10–12

43. Kozomara A, et al. miRbase: From microRNA sequences to function. Nucleic Acid Research. 2018;47:D155–D162

44. Langmean B, et al. Ultrafast and memory-efficient alignment of short DNA sequences to the human genome. Genome Biology. 2009;10:R25

45. Love MI, et al. Moderated estimation of fold change and dispersion for RNA-seq with DESeq2. Genome Biology. 2014;15:550

